# Effects of Genetically-Proxied Antihypertensive Drug Targets on Preeclampsia and Birth Weight

**DOI:** 10.64898/2026.03.17.26346752

**Authors:** Maddalena Ardissino, Alec P Morley, Eleanor M F Richards, Julia Zöllner, Buu Truong, Catherine Williamson, Michael C Honigberg, James S Ware, Kypros Nicolaides, Antonio de Marvao

**Author notes:** **Corresponding author**: Dr Antonio de Marvao School of Cardiovascular and Metabolic Medicine and Sciences, King’s College London, James Black Centre, 125 Coldharbour Lane, London SE5 9NU, UK.

## Abstract

**Background:** Preeclampsia is a leading cause of maternal and perinatal morbidity and mortality, and a major contributor to low birth weight. Beta blockers (BB) and calcium channel blockers (CCB) are the most commonly recommended agents to treat hypertension in pregnancy. Yet it remains unknown whether these agents alter the risk of preeclampsia (PE), and if so, whether effects arise through maternal physiology or through direct fetal mechanisms.

**Objectives:** To use drug-target Mendelian randomization (MR) to estimate the effects of genetically-proxied inhibition of beta-adrenergic and L-type calcium-channel targets on PE risk, birth weight, partitioned into maternal and fetal genetic components, and gestational age (GA).

**Methods:** We constructed instruments from genome-wide significant, LD-independent variants within prespecified windows around systolic blood pressure (SBP) modulating drug targets in addition to a genome-wide SBP instrument (European ancestry).

Outcomes comprised of PE (16,349 cases / 595,135 controls), maternal and fetal genetic effects on birth weight (n≈210,267 and n≈298,142), and GA (n≈151,987). Two-sample MR estimated effects per 5mmHg decrease in SBP. Bayesian colocalization assessed shared causal variants. Multiple testing was controlled with Benjamini–Hochberg correction.

**Results:** Genetically lower SBP was associated with reduced PE risk and modest increases in birth weight and GA. BB (ADRB1) target inhibition showed no convincing reduction in PE risk but was associated with lower birth weight, with associations predominantly through direct fetal genetic effects and strong colocalization at ADRB1 with fetal birth-weight signals. In contrast, CCB targets collectively associated with lower PE risk without consistent evidence of fetal growth impairment; colocalization support for individual CCB loci was limited. Sensitivity analyses (heterogeneity, pleiotropy) did not materially alter these patterns where instrument counts permitted.

**Conclusions:** Drug-target MR suggests that BB pathways are unlikely to meaningfully reduce PE and are linked to reduced fetal growth - chiefly via direct fetal mechanisms. In contrast, CCB pathways are associated with lower PE risk and largely neutral fetal growth effects. These findings support prioritizing CCBs for evaluation in comparative trials of PE prevention.

## Introduction

Preeclampsia is a leading cause of maternal and fetal morbidity and mortality^1^. The main pillars of management involve early identification of individuals at risk, initiation of preventive aspirin therapy, and strict blood pressure control once developed^2,3^.

Previous randomized clinical trials have demonstrated the efficacy of both beta blockers and calcium channel blockers for blood pressure reduction and prevention of complications^4^. However, safety concerns have been raised for beta blockers due to their association with lower birth weight^5^. Further understanding the relative efficacy and safety of these antihypertensive targets is a key priority^6^, as there are currently no adequately powered, high-quality clinical trials available to directly compare the benefits and adverse effects of the two.

In the absence of evidence from clinical trials, the Mendelian randomization (MR) framework can be used to predict safety and efficacy of pharmacological interventions. MR has previously been used to investigate the association of preeclampsia with cardiovascular disease^7–9^, its relation to genetically-predicted blood pressure traits^10^, as well as potential underlying causative circulating proteins and gene expression patterns^11,12^. In addition to this, a previous drug-target MR study identified that genetically-proxied blood pressure lowering through calcium channel blockers was associated with lower risk of preeclampsia but no change to birth weight, whereas beta blockers were not associated with reduction in preeclampsia but were associated with a reduction in birth weight^13^.

When evaluating the influence of drug target perturbation in pregnancy, it is important to bear in mind that the effects of a drug on the fetus can be attributed to two types of mechanism: indirect effects and direct effects. The former describes fetal effects that occur through the actions of the drug on maternal physiology (i.e., lowering blood pressure leading to placental hypoperfusion leading to low birth weight). The latter reflects the influence of the drug target on the fetus that occurs due to the action of the drug directly on fetal physiology (i.e., direct blockade of the drug target in the fetus leading to low birth weight). In the setting of antihypertensive therapies, both are important to consider, because both beta blockers and calcium channel blockers are known to cross the placenta, and can therefore exert direct effects on the fetus in addition to indirect effects occurring through maternal physiology. Importantly, in the previous drug-target MR study, though a safety signal for birth weight was identified^13^, a distinction was not made on whether this was related to direct or indirect effects.

In this study, we aim to leverage large-scale genetic data to evaluate the potential efficacy and safety of blood pressure lowering via beta blocker and calcium channel blocker drug targets on risk of preeclampsia and fetal birth weight, and elucidate the pathway through which potential effects on the latter might occur.

## Methods

The flowchart outlining the design of the study is depicted in **Figure 1**. This study made use of publicly available genetic association summary data from studies that sought appropriate ethical approval and participant consent. Analyses were carried out on R version 4.4.3^14^ using the TwoSampleMR (version 0.6.9)^15^, MendelianRandomization (version 0.10.0)^16^ and coloc (version 5.2.3) ^17–19^ packages.

**Figure 1.**
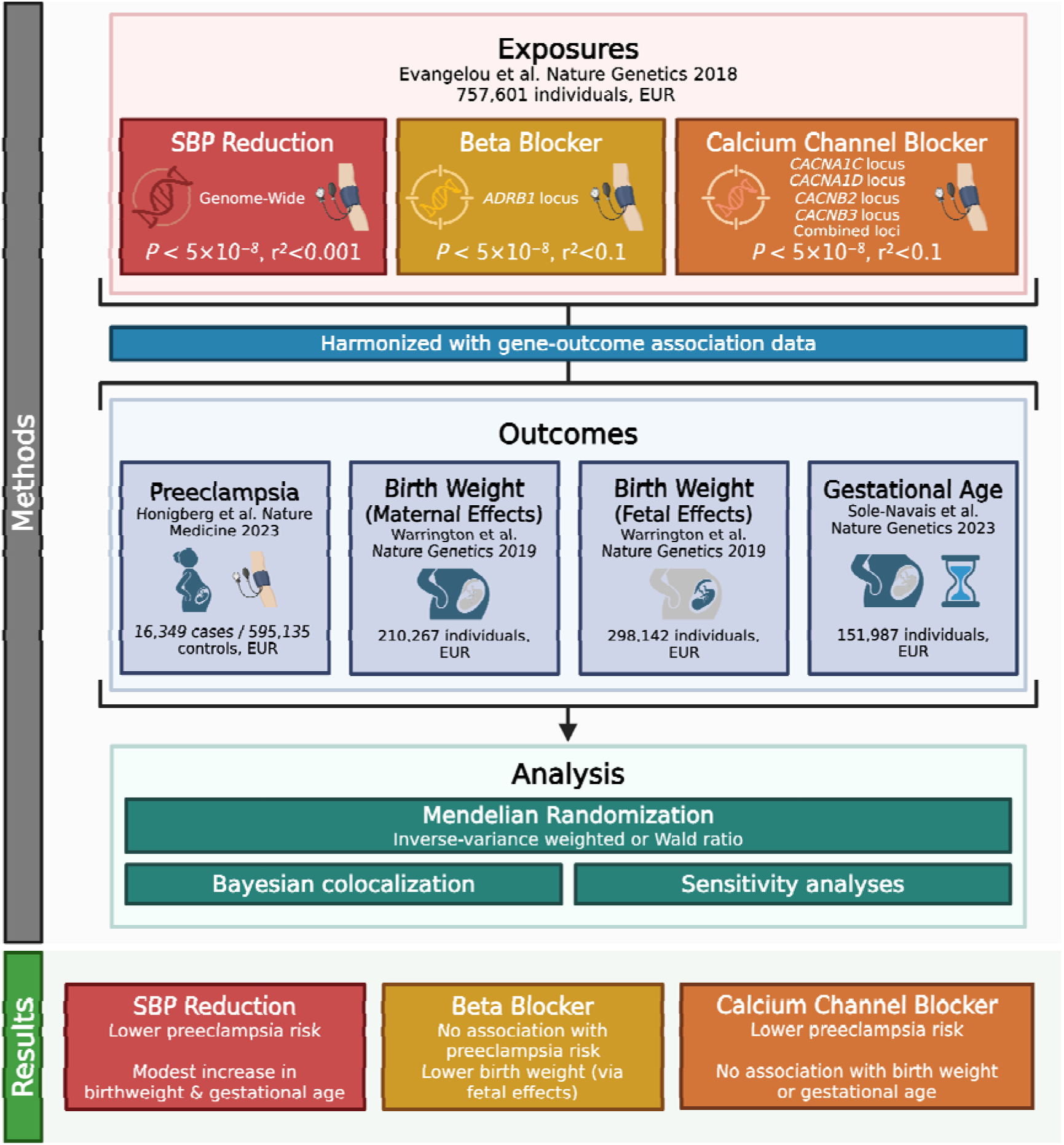
Flowchart outlining methods and key findings. EUR = European. Created in BioRender. Morley, A. (2025) https://BioRender.com/o7oup6a.

### Genetic instruments

Genome-wide significant (*P*<5×10^-8^), uncorrelated (r^2^<0.001 using the 1000 Genomes European reference panel) genetic instruments for systolic blood pressure were obtained from Evangelou et al.’s meta-analysis consisting of 757,601 individuals of European ancestry^20^. For each drug target, single-nucleotide polymorphisms in the drug target gene region (+/- 10kb) were selected with r^2^<0.1. A summary table of the data sources used in the study is available in **Supplementary Table 1**. All genetic instruments and their associations are presented in **Supplementary Table 2**.

### Outcome associations

The gene-outcome association estimates of the instruments on preeclampsia were extracted from summary data of Honigberg et al.’s GWAS on 16,349 cases with preeclampsia and 595,135 controls of European ancestry^21^. The gene-outcome association estimates of the instruments on birth weight, through both direct and indirect effects, were extracted from summary data of 210,267 individuals (indirect effects, through a maternal GWAS of child’s birth weight) and 298,142 individuals (direct effects, through a fetal GWAS of own birth weight)^22^. The gene-outcome association estimates of the instruments on gestational age were extracted from summary data of 151,987 individuals^23^.

For each analysis, SNPs that had a corresponding association estimate in the outcome GWAS were retained; unmatched SNPs were discarded, and no proxies were sought. Gene-exposure and gene-outcome association data were harmonized using the harmonize_data function in TwoSampleMR^15^. During harmonization, strand direction was inferred where possible, and palindromic SNPs were harmonized. If this was not possible due to incompatible or ambiguous alleles, the SNP was excluded from further analysis.

### Statistical analysis

Inverse variance-weighted MR was performed to estimate the association between the systolic blood pressure lowering overall and through specific drug target and outcome, for instrumental variable sets with more than one variant^24,25^. Where only one instrument was present, the Wald ratio^26^ method was used. Results for the primary analyses are presented as odds ratios (OR) and 95% confidence intervals (95%CI) for every 5mmHg reduction in systolic blood pressure for preeclampsia, and beta coefficients (β) and 95% confidence intervals (95%CI) for birth weight and gestational age. The results of the main analyses were corrected for using the Benjamini-Hochberg correction of *P*-values, with an expected 5% false discovery rate (FDR).

### Sensitivity analyses

For the results of MR studies to be valid, instruments must satisfy three key instrumental variable assumptions^27^:

1. Relevance: the variants are associated with the exposure
2. Independence: there are no common causes of the genetic variant, which largely refers to population stratification, a phenomenon of confounding which can occur when populations of different ancestry are used in the same MR analysis
3. Exclusion restriction: the only pathway through which the variant influences the outcome is through the exposure

The first assumption can be addressed through calculation of the instrument F-statistics, which we performed using the formula:

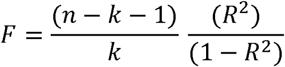

where R^2^ is the explained variance in the regression of all SNPs, *n* is the number of participants in the study, *k* is the number of instrumental variants. The R^2^ was calculated as the sum of SNP-wise R^2^ of instruments, which is obtained as follows:

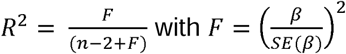

where *β* represents the effect size of the genetic variant in the exposure GWAS, and

*SE*(*β*) represents the standard error of the effect size of the genetic variant in the exposure GWAS. Broadly, F-statistics > 10 reassure against the presence of weak instrumental bias.

The second assumption relating to population stratification, was limited through selection of data sources for both gene-exposure and gene-outcome associations that (1) adjusted for genetic principal components and (2) included only European ancestry populations.

The third assumption cannot be formally tested for, but can be evaluated in a multi-layer approach through sensitivity analyses, using MR-Egger and weighted median MR, which we performed where possible (when ≥3 instrumental SNPs were available)^28^.

### Bayesian colocalization analyses

For any genetically-predicted exposure and outcome pair, Bayesian colocalization analyses can be used to support the results of MR analyses^17^. Briefly, this method evaluates the likelihood that the same variants are causal for both the exposure and the outcome, which strengthens the evidence to support a causal association between the two. Specifically, Bayesian colocalization assess the posterior probability of genetic variants within a specific gene region having:

- No association with either exposure or outcome (PP.H0)
- Association with exposure, not with outcome (PP.H1)
- Association with outcome, not with exposure (PP.H2)
- Association with exposure and outcome, two distinct causal SNPs (PP.H3)
- Association with exposure and outcome, both due to a single causal SNP (PP.H4)

Where PP.H4 of ≥80% can be considered strong evidence of colocalization. In addition to this, the PP.H4 / (PP.H3+PP.H4) value was calculated. The value PP.H4 / (PP.H3+PP.H4) essentially denotes the probability of colocalization conditional on the presence of a causal variant for the outcome. In cases when the PP.H1 or PP.H2 are high, this can help to distinguish whether the remaining evidence tilts in the favor of colocalization or non-colocalization.

For the colocalization analysis, genetic variants in the drug target gene regions were extracted in the systolic blood pressure GWAS. Variants in the corresponding position were then extracted from the outcome GWAS data. Colocalization analyses were performed using coloc R package v5.2.3^17–19^ with prior probabilities set as p_1_=1×10^-4^, p_2_=1×10^-4^, and p_12_=1×10^-5^.

## Results

Genetically-proxied systolic blood pressure reduction was associated with markedly lower risk of preeclampsia (OR 0.76 [0.75 to 0.78] *P* = 2.74×10⁻¹□□), higher birth weight via indirect maternal effects (β 0.06 [0.05 to 0.07] *P* = 4.93×10⁻¹□), higher birth weight via direct fetal effects (β 0.05 [0.04 to 0.05] *P* = 6.41×10⁻□□), and longer gestational age (β 0.22 [0.15 to 0.29] *P* = 1.66×10⁻□).

Genetically-proxied blood pressure lowering via calcium channel blockade, representing combined effects across CACNA1C, CACNA1D, CACNB2, and CACNB3 drug targets, was associated with lower risk of preeclampsia (OR 0.77 [0.69 to 0.86] *P* = 8.02×10⁻□, probability of shared causal variant [PP.H4]: 1.7–5.9%, PP.H4/(PP.H3+PP.H4): 30.2–97.1%), but not with birth weight via either maternal or fetal effects, or gestational age.

Among individual CCB targets, CACNA1C inhibition was associated with higher birth weight via direct fetal effects (β 0.15 [0.06 to 0.25] *P* = 5.99×10⁻³, PP.H4: 1.1%, PP.H4/(PP.H3+PP.H4): 33.5%), but had no statistically significant association with gestational age (β −1.57 [−3.14 to 0.01] *P* = 0.130, PP.H4: 1.0%, PP.H4/(PP.H3+PP.H4): 39.0%). CACNA1D inhibition showed no association with birth weight via either maternal or fetal effects, gestational age or risk of preeclampsia. CACNB2 inhibition was associated with lower risk of preeclampsia (OR 0.71 [0.63 to 0.80] *P* = 2.78×10⁻□, PP.H4: 5.9%, PP.H4/(PP.H3+PP.H4): 51.1%) and a trend towards lower birth weight via direct fetal effects (β −0.03 [−0.05 to 0.00] *P* = 0.109, PP.H4: 0.6%, PP.H4/(PP.H3+PP.H4): 15.6%) but had no associations with birth weight via maternal effects, or gestational age. CACNB3 inhibition showed no significant associations with birth weight, gestational age, or preeclampsia.

Genetically-proxied ADRB1 inhibition, proxying beta blocker drug targets, was associated with lower birth weight via both indirect maternal effects (β −0.23 [−0.30 to −0.16] *P* = 1.41×10⁻¹□, PP.H4: 100%, PP.H4/(PP.H3+PP.H4): 100%) and direct fetal effects (β −0.30 [−0.43 to −0.18] *P* = 8.88×10⁻□, PP.H4: 99.0%, PP.H4/(PP.H3+PP.H4): 99.0%), but not with gestational age (β −0.36 [−1.45 to 0.72] *P* = 0.553, PP.H4: 0.4%, PP.H4/(PP.H3+PP.H4): 93.7%) or preeclampsia (OR 0.62 [0.25 to 1.51] *P* = 0.408, PP.H4: 98.1%, PP.H4/(PP.H3+PP.H4): 98.9%).

The results of the main analyses are summarized in **Figure 2**, and colocalization results are reported in **Figure 3**. All instrument F-statistics were greater than 10, as reported in **Supplementary Table 3**. There was no evidence suggestive of significant pleiotropy in the sensitivity analyses which would materially impact estimates, the results of which are reported in **Supplementary Table 4**.

**Figure 2.**
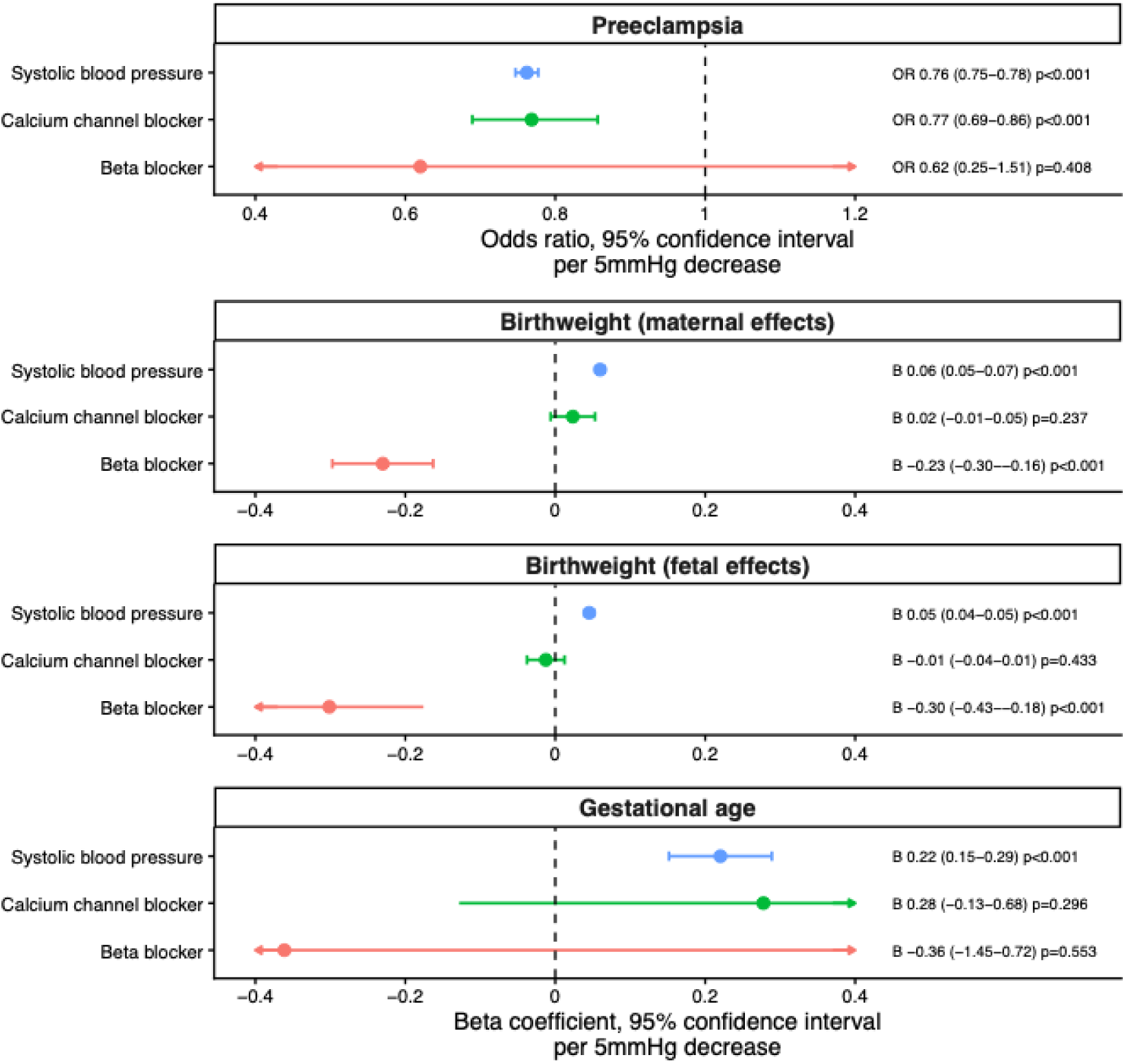
Genetic associations of systolic blood pressure lowering overall and via beta blocker and calcium channel blocker drug targets on pregnancy outcomes. OR = odds ratio.

**Figure 3.**
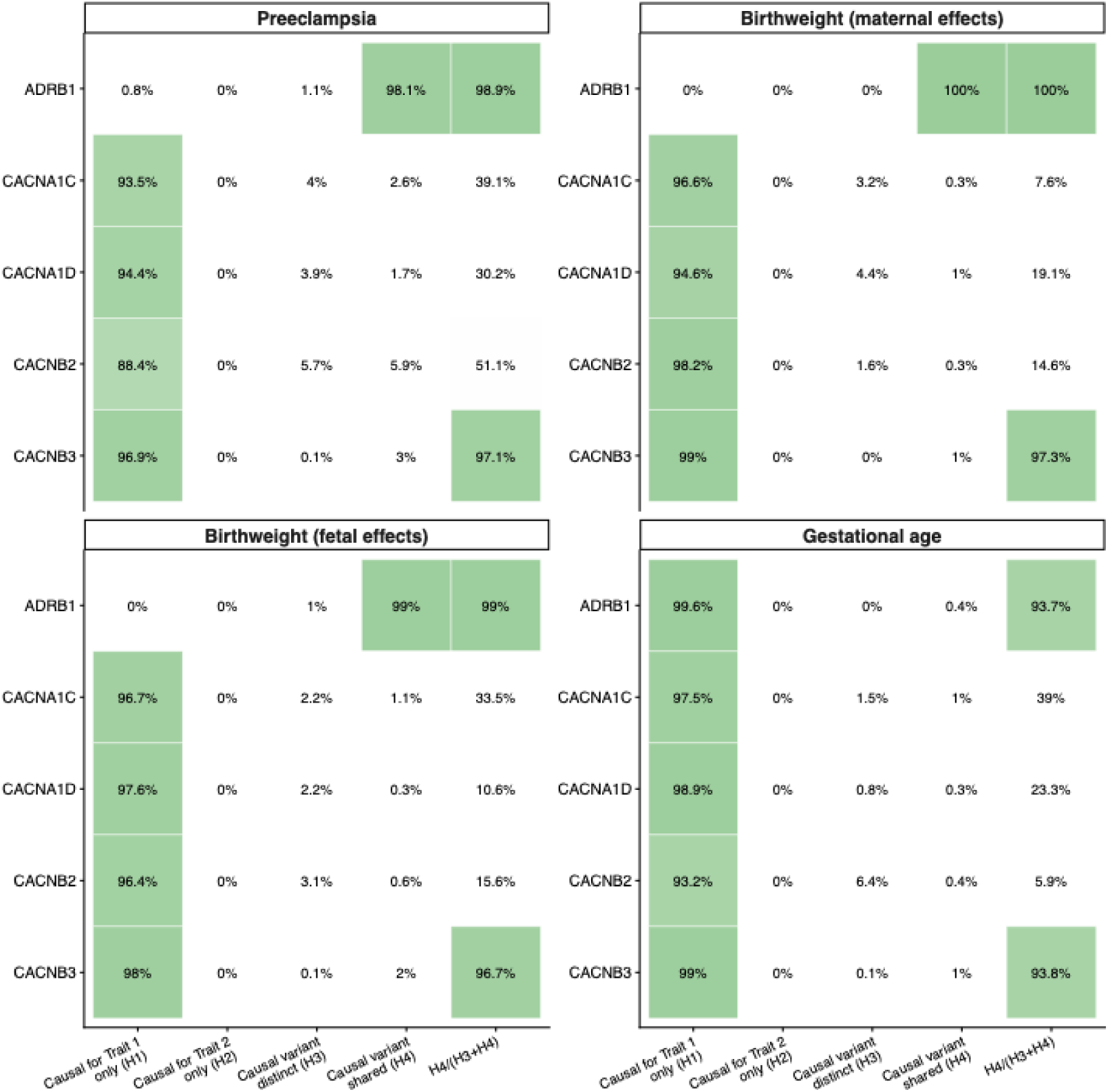
Results of Bayesian colocalization analysis for the probability of shared causal variants within drug target genes with pregnancy outcomes.

**Table 1.** Associations of genetically-proxied systolic blood pressure reduction (by 5mmHg) overall, by beta blockade and by calcium channel blockade on outcome of preeclampsia, birth weight (via indirect maternal and direct fetal effects), and gestational age. CI = Confidence interval.

| **Preeclampsia** |  |  |  |  |  |  |
| --- | --- | --- | --- | --- | --- | --- |
|  | **Exposure** | **Odds Ratio** | **Lower 95%-CI** | **Upper 95%-CI** | **P-value** | **P-value (adjusted)** |
| | Systolic blood pressure | 0.762 | 0.747 | 0.777 | $9.80 \times 10^{-158}$ | $2.74 \times 10^{-156}$ |
| | Calcium channel blocker (Overall) | 0.768 | 0.689 | 0.857 | $2.01 \times 10^{-6}$ | $8.02 \times 10^{-6}$ |
|  | Calcium channel blocker (CACNA1C) | 1.395 | 0.873 | 2.229 | 0.163 | 0.286 |
|  | Calcium channel blocker (CACNA1D) | 0.806 | 0.615 | 1.056 | 0.118 | 0.237 |
| | Calcium channel blocker (CACNB2) | 0.711 | 0.628 | 0.804 | $5.97 \times 10^{-8}$ | $2.78 \times 10^{-7}$ |
|  | Calcium channel blocker (CACNB3) | 1.217 | 0.730 | 2.028 | 0.452 | 0.527 |
|  | Beta blocker (ADRB1) | 0.620 | 0.255 | 1.507 | 0.292 | 0.408 |
| **Birth weight (maternal effects)** |  |  |  |  |  |  |
|  | **Exposure** | **Beta coefficient** | **Lower 95%-CI** | **Upper 95%-CI** | **P-value** | **P-value (adjusted)** |
| | Systolic blood pressure | 0.060 | 0.054 | 0.066 | $3.52 \times 10^{-82}$ | $4.93 \times 10^{-81}$ |
|  | Calcium channel blocker (Overall) | 0.023 | -0.006 | 0.053 | 0.119 | 0.237 |
|  | Calcium channel blocker (CACNA1C) | 0.031 | -0.083 | 0.145 | 0.595 | 0.617 |
|  | Calcium channel blocker (CACNA1D) | 0.056 | -0.098 | 0.209 | 0.475 | 0.532 |
|  | Calcium channel blocker (CACNB2) | 0.016 | -0.013 | 0.044 | 0.282 | 0.408 |
|  | Calcium channel blocker (CACNB3) | 0.046 | -0.068 | 0.160 | 0.426 | 0.518 |
| | Beta blocker (ADRB1) | -0.230 | -0.297 | -0.163 | $2.01 \times 10^{-11}$ | $1.41 \times 10^{-10}$ |
| **Birth weight (fetal effects)** |  |  |  |  |  |  |
|  | **Exposure** | **Beta coefficient** | **Lower 95%-CI** | **Upper 95%-CI** | **P-value** | **P-value (adjusted)** |
| | Systolic blood pressure | 0.045 | 0.039 | 0.051 | $6.86 \times 10^{-51}$ | $6.41 \times 10^{-50}$ |
|  | Calcium channel blocker (Overall) | -0.013 | -0.038 | 0.012 | 0.325 | 0.433 |
| | Calcium channel blocker (CACNA1C) | 0.152 | 0.056 | 0.247 | $1.93 \times 10^{-3}$ | $5.99 \times 10^{-3}$ |
|  | Calcium channel blocker (CACNA1D) | -0.029 | -0.096 | 0.039 | 0.410 | 0.518 |
| | Calcium channel blocker (CACNB2) | -0.025 | -0.050 | -0.001 | $3.88 \times 10^{-2}$ | 0.109 |
|  | Calcium channel blocker (CACNB3) | 0.076 | -0.022 | 0.173 | 0.127 | 0.237 |
| | Beta blocker (ADRB1) | -0.301 | -0.427 | -0.176 | $2.54 \times 10^{-6}$ | $8.88 \times 10^{-6}$ |
| **Gestational age** |  |  |  |  |  |  |
|  | **Exposure** | **Beta coefficient** | **Lower 95%-CI** | **Upper 95%-CI** | **P-value** | **P-value (adjusted)** |
| | Systolic blood pressure | 0.220 | 0.152 | 0.289 | $2.97 \times 10^{-10}$ | $1.66 \times 10^{-9}$ |
|  | Calcium channel blocker (Overall) | 0.278 | -0.128 | 0.684 | 0.180 | 0.296 |
|  | Calcium channel blocker (CACNA1C) | -1.567 | -3.143 | 0.009 | 0.051 | 0.130 |
|  | Calcium channel blocker (CACNA1D) | 0.679 | -0.568 | 1.927 | 0.286 | 0.408 |
|  | Calcium channel blocker (CACNB2) | 0.353 | -0.097 | 0.802 | 0.124 | 0.237 |
|  | Calcium channel blocker (CACNB3) | 0.244 | -1.778 | 2.266 | 0.813 | 0.813 |
|  | Beta blocker (ADRB1) | -0.361 | -1.446 | 0.723 | 0.514 | 0.553 |

## Discussion

In this study, we show that genetically-proxied reductions in systolic blood pressure are associated with lower risk of preeclampsia, higher birth weight, and longer gestational age, as previously demonstrated in both observational and genetic studies^7–11,29^. However, these associations differed by antihypertensive drug target. Genetic proxies for beta blocker targets (ADRB1), showed no evidence of reduced preeclampsia risk, but were associated with lower birth weight, driven predominantly by direct fetal effects and to a lesser extent by indirect maternal effects. In contrast, calcium channel blocker targets (CACNA1C, CACNA1D, CACNB2, CACNB3) showed a protective association with preeclampsia without evidence of adverse effects on fetal growth.

We have previously shown that genetically-proxied beta blockade was associated with reduced fetal growth, using Mendelian randomization in a different, smaller preeclampsia cohort^13^. Furthermore, other studies have investigated the effect of genetically-proxied antihypertensives on composite hypertensive disorders of pregnancy, however, did not investigate effects on fetal growth, nor preeclampsia specifically^30^. Observational studies have similarly found that use of beta blockers in pregnancy is associated with small for gestational age (SGA) babies^31,32^ such as a Danish study of 911,685 births including 2,459 pregnancies exposed to beta blockers reporting almost double the odds of SGA^5^. However, a Cochrane systematic review of antihypertensive therapy in pregnancy for women with mild to moderate hypertension found that there is no evidence of a difference in the risk of SGA when methyldopa and calcium channel blockers together are compared with beta blockers although only one of these studies compared calcium channel blockers to beta blockers^4^. As such, the Cochrane review concludes that high-quality, large randomized controlled trials are still required.

Building on the existing literature, we further explored the mechanistic origin of these effects by disentangling maternal and fetal pathways. Colocalization analyses at the ADRB1 locus demonstrated a very high probability of shared causal variants in both fetal and maternal pathways, supporting a contribution from direct fetal genetic effects alongside indirect maternal influences. These findings, support the hypothesis that beta blocker exposure may reduce fetal cardiac output or alter metabolic function after transplacental transfer. This raises the possibility that such adverse effects might be attenuated through the development or preferential use of beta blockers with reduced placental transfer, a hypothesis that warrants further experimental and clinical investigation.

By contrast, blood pressure lowering via calcium channel blockade was consistently associated with a lower risk of preeclampsia, without consistent evidence of adverse effects on fetal growth. Although most associations with maternal birth weight effects were not statistically significant, they were directionally positive – suggesting that blood pressure lowering through this pathway may even support improved fetal growth. While one individual target (CACNB2) showed evidence of lower fetal birth weight, this finding was not consistent across other CCB targets, arguing against a class-wide effect. Colocalization evidence was modest, reflecting limited power at some loci, but the PP.H4/(PP.H3+PP.H4) values were generally reasonable, cautiously supporting shared causal relevance.

The Cochrane review of antihypertensive therapy in pregnancy for women with mild to moderate hypertension found no significant difference in risk of severe hypertension when comparing calcium channel blockers to beta blockers, but again only one relevant randomized trial was identified^33^. For the outcome of proteinuria or preeclampsia, two trials with 204 women compared calcium channel blockers to beta blockers and found no significant difference, however a subsequent systematic review and network meta-analysis found that labetalol decreased proteinuria/ preeclampsia when compared with calcium channel blockers^33^. These conflicting results reflect a paucity of high-quality trial data, although a randomized trial of nifedipine versus labetalol in women with hypertension in pregnancy in the UK successfully recruited 2,254 participants by January 2025 and the results of this study are eagerly awaited^34^.

Current NICE guidelines for hypertension in pregnancy recommend labetalol as first-line and nifedipine as second-line treatments^35^. In the postnatal setting, calcium channel blockers are recommended first-line therapy for women of African or Caribbean ancestry, reflecting broader adult hypertension guidance and recognized differences in hypertension etiology by ethnicity. The present analyses were limited by reliance on genetic data predominantly from individuals of European ancestry.

Therefore, extrapolation to other ancestral populations should be undertaken with caution, and replication in more diverse cohorts is warranted. When interpreting these results, it is also important to recognize that Mendelian randomization estimates should not be used to infer effect size of antihypertensive therapy in pregnancy, but rather to indicate the presence and direction of causal effects. The genetic effect sizes reflect small, lifelong perturbations of the drug target, whereas pharmacologic treatments induce larger and temporally restricted physiological changes.

An important conceptual distinction should also be drawn between the preeclampsia phenotype examined in this study, reflecting the incidence *or* risk of developing the condition, and the clinical management of established preeclampsia. While genetically-proxied inhibition of beta blocker targets did not associate with a lower risk of preeclampsia incidence, this should not be interpreted as evidence that beta blockers are ineffective for blood pressure control or prevention of complications in women with preeclampsia. Our findings pertain specifically to disease susceptibility and causal pathways leading to preeclampsia onset, rather than therapeutic efficacy in managing preeclampsia once it has developed. This distinction is critical, as antihypertensive efficacy and safety profiles in the setting of established disease may differ substantially from their influence on disease risk. Conversely, the protective associations observed for calcium channel blocker targets raise the important question of whether these mechanisms could be explored further for primary prevention of preeclampsia, particularly if initiated early in pregnancy among women at high risk, potentially in combination with aspirin therapy.

In summary, our results suggest that beta blocker mediated blood pressure lowering is unlikely to reduce preeclampsia risk and is associated with lower birth weight, primarily through direct fetal effects. In contrast, blood pressure lowering via calcium channel blocker pathways was associated with reduced pre-eclampsia risk and potentially favorable for fetal growth. Together, these findings underscore the need for randomized controlled trials, alongside mechanistic studies, to evaluate whether calcium channel blocker based antihypertensive strategies can be leveraged for the prevention of pre-eclampsia and the optimization of perinatal outcomes.

## Supporting information

STROBE-MR checklist

## Data Availability

All summary-level genetic association data used in this study are publicly available from the original genome-wide association studies cited in the manuscript. No individual-level data were used in this study. All data sources are referenced in the manuscript, and access details are provided in the Supplementary Material.

## Acknowledgements

The authors acknowledge all investigators and participants of the studies contributing to the present analyses.

The Graphical Abstract was created in BioRender. Morley, A. (2026) https://BioRender.com/o7oup6a. License to use the BioRender content, including icons, templates and other original artwork was granted to Alec P Morley (Agreement number YH29DITMLK).

## Data availability statement

- The genetic analyses in this study utilize summary-level datasets which are available from the cited sources.

## Ethical approval

- Ethical approval was not required for the genetic analysis. The genetic analysis used publicly available genome-wide association study (GWAS) summary data available to download at cited sources. Ethical approval and participant consent were obtained in the original publications.

