## Supplementary material for "Effects of Genetically-Proxied Antihypertensive Drug Targets on Preeclampsia and Birth Weight": STROBE-MR checklist

**STROBE-MR checklist of recommended items to address in reports of Mendelian randomization studies**^1^ ^2^

| **Item No.** | **Section** | **Checklist item** | **Page No.** | **Relevant text from manuscript** |
| --- | --- | --- | --- | --- |
| 1 | **TITLE and ABSTRACT** | Indicate Mendelian randomization (MR) as the study’s design in the title and/or the abstract if that is a main purpose of the study | 3 | *“To use drug-target Mendelian randomization (MR) to estimate the effects of genetically-proxied inhibition of beta-adrenergic and L-type calcium-channel targets on PE risk, birth weight, partitioned into maternal and fetal genetic components, and gestational age (GA).”* |
|  | **INTRODUCTION** |  |  |  |
| 2 | **Background** | Explain the scientific background and rationale for the reported study. What is the exposure? Is a potential causal relationship between exposure and outcome plausible? Justify why MR is a helpful method to address the study question | 5 | *“In the absence of evidence from clinical trials, the Mendelian randomization (MR) framework can be used to predict safety and efficacy of pharmacological interventions. MR has previously been used to investigate the association of preeclampsia with cardiovascular disease7–9, its relation to genetically-predicted blood pressure traits10, as well as potential underlying causative circulating proteins and gene expression patterns11,12. In addition to this, a previous drug-target MR study identified that genetically-proxied blood pressure lowering through calcium channel blockers was associated with lower risk of preeclampsia but no change to birth weight, whereas beta blockers were not associated with reduction in preeclampsia but were associated with a reduction in birth weight13.”* |
| 3 | **Objectives** | State specific objectives clearly, including pre-specified causal hypotheses (if any). State that MR is a method that, under specific assumptions, intends to estimate causal effects | 6 | *“In this study, we aim to leverage large-scale genetic data to evaluate the potential efficacy and safety of blood pressure lowering via beta blocker and calcium channel blocker drug targets on risk of preeclampsia and fetal birth weight, and elucidate the pathway through which potential effects on the latter might occur.”* |
|  | **METHODS** |  |  |  |
| 4 | **Study design and data sources** | Present key elements of the study design early in the article. Consider including a table listing sources of data for all phases of the study. For each data source contributing to the analysis, describe the following: |  |  |
|  | a) | Setting: Describe the study design and the underlying population, if possible. Describe the setting, locations, and relevant dates, including periods of recruitment, exposure, follow-up, and data collection, when available. | 7  N/A  N/A | Methods  Figure 1  Supplementary Table 1 – publicly available GWAS summary statistics |
|  | b) | Participants: Give the eligibility criteria, and the sources and methods of selection of participants. Report the sample size, and whether any power or sample size calculations were carried out prior to the main analysis | 7  N/A  N/A | Methods  Figure 1  Supplementary Table 1 – publicly available GWAS summary statistics |
|  | c) | Describe measurement, quality control and selection of genetic variants | 7  N/A  N/A | Methods  Figure 1  Supplementary Table 1 – publicly available GWAS summary statistics |
|  | d) | For each exposure, outcome, and other relevant variables, describe methods of assessment and diagnostic criteria for diseases | N/A | Supplementary Table 1 – summary table of data sources |
|  | e) | Provide details of ethics committee approval and participant informed consent, if relevant | 7 | *“This study made use of publicly available genetic association summary data from studies that sought appropriate ethical approval and participant consent.”* |
| 5 | **Assumptions** | Explicitly state the three core IV assumptions for the main analysis (relevance, independence and exclusion restriction) as well assumptions for any additional or sensitivity analysis | 8-9 | *“For the results of MR studies to be valid, instruments must satisfy three key instrumental variable assumptions27:*  *(1) Relevance: the variants are associated with the exposure*  *(2) Independence: there are no common causes of the genetic variant, which largely refers to population stratification, a phenomenon of confounding which can occur when populations of different ancestry are used in the same MR analysis*  *(3) Exclusion restriction: the only pathway through which the variant influences the outcome is through the exposure”* |
| 6 | **Statistical methods: main analysis** | Describe statistical methods and statistics used |  |  |
|  | a) | Describe how quantitative variables were handled in the analyses (i.e., scale, units, model) | 8 | *“Results for the primary analyses are presented as odds ratios (OR) and 95% confidence intervals (95%CI) for every 5mmHg reduction in systolic blood pressure for preeclampsia, and beta coefficients (β) and 95% confidence intervals (95%CI) for birth weight and gestational age.”* |
|  | b) | Describe how genetic variants were handled in the analyses and, if applicable, how their weights were selected | 8 | *“For each analysis, SNPs that had a corresponding association estimate in the outcome GWAS were retained; unmatched SNPs were discarded, and no proxies were sought. Gene-exposure and gene-outcome association data were harmonized using the harmonize_data function in TwoSampleMR15. During harmonization, strand direction was inferred where possible, and palindromic SNPs were harmonized. If this was not possible due to incompatible or ambiguous alleles, the SNP was excluded from further analysis…. Results for the primary analyses are presented as odds ratios (OR) and 95% confidence intervals (95%CI) for every 5mmHg reduction in systolic blood pressure for preeclampsia, and beta coefficients (β) and 95% confidence intervals (95%CI) for birth weight and gestational age.”* |
|  | c) | Describe the MR estimator (e.g. two-stage least squares, Wald ratio) and related statistics. Detail the included covariates and, in case of two-sample MR, whether the same covariate set was used for adjustment in the two samples | 8 | *“Inverse variance-weighted MR was performed to estimate the association between the systolic blood pressure lowering overall and through specific drug target and outcome, for instrumental variable sets with more than one variant24,25. Where only one instrument was present, the Wald ratio26 method was used”* |
|  | d) | Explain how missing data were addressed | 8 | *“For each analysis, SNPs that had a corresponding association estimate in the outcome GWAS were retained; unmatched SNPs were discarded, and no proxies were sought.”* |
|  | e) | If applicable, indicate how multiple testing was addressed | 8 | *“The results of the main analyses were corrected for using the Benjamini-Hochberg correction of P-values, with an expected 5% false discovery rate (FDR).”* |
| 7 | **Assessment of assumptions** | Describe any methods or prior knowledge used to assess the assumptions or justify their validity | 9 | *“(3) Exclusion restriction: the only pathway through which the variant influences the outcome is through the exposure*  *The first assumption can be addressed through calculation of the instrument F-statistics, which we performed using the formula:*  *F=((n-k-1))/k ((R^2))/((1-R^2))*  *where R^2 is the explained variance in the regression of all SNPs, n is the number of participants in the study, k is the number of instrumental variants. The R^2 was calculated as the sum of SNP-wise R^2 of instruments, which is obtained as follows:*  *R^2= F/((n-2+F)) with F=(β/(SE(β)))^2*  *where  represents the effect size of the genetic variant in the exposure GWAS, and SE( ) represents the standard error of the effect size of the genetic variant in the exposure GWAS. Broadly, F-statistics > 10 reassure against the presence of weak instrumental bias.*  *The second assumption relating to population stratification, was limited through selection of data sources for both gene-exposure and gene-outcome associations that (1) adjusted for genetic principal components and (2) included only European ancestry populations.*  *The third assumption cannot be formally tested for, but can be evaluated in a multi-layer approach through sensitivity analyses, using MR-Egger and weighted median MR, which we performed where possible (when ≥3 instrumental SNPs were available)28.”* |
| 8 | **Sensitivity analyses and additional analyses** | Describe any sensitivity analyses or additional analyses performed (e.g. comparison of effect estimates from different approaches, independent replication, bias analytic techniques, validation of instruments, simulations) | 9-10 | *“Bayesian colocalization analyses*  *For any genetically-predicted exposure and outcome pair, Bayesian colocalization analyses can be used to support the results of MR analyses17. Briefly, this method evaluates the likelihood that the same variants are causal for both the exposure and the outcome, which strengthens the evidence to support a causal association between the two. Specifically, Bayesian colocalization assess the posterior probability of genetic variants within a specific gene region having…”* |
| 9 | **Software and pre-registration** |  |  |  |
|  | a) | Name statistical software and package(s), including version and settings used | 7 | *“Analyses were carried out on R version 4.4.314 using the TwoSampleMR (version 0.6.9)15, MendelianRandomization (version 0.10.0)16 and coloc (version 5.2.3) 17–19packages.”* |
|  | b) | State whether the study protocol and details were pre-registered (as well as when and where) | 7 | N/A use of publicly available GWAS summary statistics |
|  | **RESULTS** |  |  |  |
| 10 | **Descriptive data** |  |  |  |
|  | a) | Report the numbers of individuals at each stage of included studies and reasons for exclusion. Consider use of a flow diagram | 23 | Figure 1 |
|  | b) | Report summary statistics for phenotypic exposure(s), outcome(s), and other relevant variables (e.g. means, SDs, proportions) | N/A | Supplementary Table 2 |
|  | c) | If the data sources include meta-analyses of previous studies, provide the assessments of heterogeneity across these studies | N/A | N/A |
|  | d) | For two-sample MR:  i.  Provide justification of the similarity of the genetic variant-exposure associations between the exposure and outcome samples  ii.  Provide information on the number of individuals who overlap between the exposure and outcome studies | N/A | Different data sets |
| 11 | **Main results** |  |  |  |
|  | a) | Report the associations between genetic variant and exposure, and between genetic variant and outcome, preferably on an interpretable scale | N/A | Supplementary Table 2 |
|  | b) | Report MR estimates of the relationship between exposure and outcome, and the measures of uncertainty from the MR analysis, on an interpretable scale, such as odds ratio or relative risk per SD difference | 11  24  26-27 | Results  Figure 2  Table 1 |
|  | c) | If relevant, consider translating estimates of relative risk into absolute risk for a meaningful time period | N/A | N/A |
|  | d) | Consider plots to visualize results (e.g. forest plot, scatterplot of associations between genetic variants and outcome versus between genetic variants and exposure) | 24 | Figure 2 |
| 12 | **Assessment of assumptions** |  |  |  |
|  | a) | Report the assessment of the validity of the assumptions | N/A | Supplementary Table 3 – F statistics  Supplementary Table 4 – Sensitivity Analyses |
|  | b) | Report any additional statistics (e.g., assessments of heterogeneity across genetic variants, such as *I^2^*, Q statistic or E-value) | N/A |  |
| 13 | **Sensitivity analyses and additional analyses** |  |  |  |
|  | a) | Report any sensitivity analyses to assess the robustness of the main results to violations of the assumptions | N/A | Supplementary Table 3 – F statistics  Supplementary Table 4 – Sensitivity Analyses |
|  | b) | Report results from other sensitivity analyses or additional analyses | 11-12  25 | Results  Figure 3 – Colocalization results |
|  | c) | Report any assessment of direction of causal relationship (e.g., bidirectional MR) | N/A | N/A |
|  | d) | When relevant, report and compare with estimates from non-MR analyses | N/A | N/A |
|  | e) | Consider additional plots to visualize results (e.g., leave-one-out analyses) | N/A | N/A |
|  | **DISCUSSION** |  |  |  |
| 14 | **Key results** | Summarize key results with reference to study objectives | 13 | *“In this study, we show that genetically-proxied reductions in systolic blood pressure are associated with lower risk of preeclampsia, higher birth weight, and longer gestational age, as previously demonstrated in both observational and genetic studies7–11,29. However, these associations differed by antihypertensive drug target. Genetic proxies for beta blocker targets (ADRB1), showed no evidence of reduced preeclampsia risk, but were associated with lower birth weight, driven predominantly by direct fetal effects and to a lesser extent by indirect maternal effects. In contrast, calcium channel blocker targets (CACNA1C, CACNA1D, CACNB2, CACNB3) showed a protective association with preeclampsia without evidence of adverse effects on fetal growth.”* |
| 15 | **Limitations** | Discuss limitations of the study, taking into account the validity of the IV assumptions, other sources of potential bias, and imprecision. Discuss both direction and magnitude of any potential bias and any efforts to address them |  |  |
| 16 | **Interpretation** |  |  |  |
|  | a) | Meaning: Give a cautious overall interpretation of results in the context of their limitations and in comparison with other studies | 15-16 | *“An important conceptual distinction should also be drawn between the preeclampsia phenotype examined in this study, reflecting the incidence or risk of developing the condition, and the clinical management of established preeclampsia. While genetically-proxied inhibition of beta blocker targets did not associate with a lower risk of preeclampsia incidence, this should not be interpreted as evidence that beta blockers are ineffective for blood pressure control or prevention of complications in women with preeclampsia. Our findings pertain specifically to disease susceptibility and causal pathways leading to preeclampsia onset, rather than therapeutic efficacy in managing preeclampsia once it has developed. This distinction is critical, as antihypertensive efficacy and safety profiles in the setting of established disease may differ substantially from their influence on disease risk. Conversely, the protective associations observed for calcium channel blocker targets raise the important question of whether these mechanisms could be explored further for primary prevention of preeclampsia, particularly if initiated early in pregnancy among women at high risk, potentially in combination with aspirin therapy.”* |
|  | b) | Mechanism: Discuss underlying biological mechanisms that could drive a potential causal relationship between the investigated exposure and the outcome, and whether the gene-environment equivalence assumption is reasonable. Use causal language carefully, clarifying that IV estimates may provide causal effects only under certain assumptions | 12-16 | Discussion |
|  | c) | Clinical relevance: Discuss whether the results have clinical or public policy relevance, and to what extent they inform effect sizes of possible interventions | 16 | *“In summary, our results suggest that beta blocker mediated blood pressure lowering is unlikely to reduce preeclampsia risk and is associated with lower birth weight, primarily through direct fetal effects. In contrast, blood pressure lowering via calcium channel blocker pathways was associated with reduced pre-eclampsia risk and potentially favorable for fetal growth. Together, these findings underscore the need for randomized controlled trials, alongside mechanistic studies, to evaluate whether calcium channel blocker based antihypertensive strategies can be leveraged for the prevention of pre-eclampsia and the optimization of perinatal outcomes.”* |
| 17 | **Generalizability** | Discuss the generalizability of the study results (a) to other populations, (b) across other exposure periods/timings, and (c) across other levels of exposure | 15-16 | *“An important conceptual distinction should also be drawn between the preeclampsia phenotype examined in this study, reflecting the incidence or risk of developing the condition, and the clinical management of established preeclampsia. While genetically-proxied inhibition of beta blocker targets did not associate with a lower risk of preeclampsia incidence, this should not be interpreted as evidence that beta blockers are ineffective for blood pressure control or prevention of complications in women with preeclampsia. Our findings pertain specifically to disease susceptibility and causal pathways leading to preeclampsia onset, rather than therapeutic efficacy in managing preeclampsia once it has developed. This distinction is critical, as antihypertensive efficacy and safety profiles in the setting of established disease may differ substantially from their influence on disease risk. Conversely, the protective associations observed for calcium channel blocker targets raise the important question of whether these mechanisms could be explored further for primary prevention of preeclampsia, particularly if initiated early in pregnancy among women at high risk, potentially in combination with aspirin therapy.”* |
|  | **OTHER INFORMATION** |  |  |  |
| 18 | **Funding** | Describe sources of funding and the role of funders in the present study and, if applicable, sources of funding for the databases and original study or studies on which the present study is based | 28 | *“MA is supported by a Medical Research Council Clinical Research Training Fellowship (MR/Z505146/1). MCH is supported by the U.S. National Heart, Lung, and Blood Institute (NHLBI, R01HL173028) and the American Heart Association (24RGRSG1275749, 25SFRNCCKMS1443062, 25SFRNPCKMS1463898). AdM is supported by the Fetal Medicine Foundation (495237).”* |
| 19 | **Data and data sharing** | Provide the data used to perform all analyses or report where and how the data can be accessed, and reference these sources in the article. Provide the statistical code needed to reproduce the results in the article, or report whether the code is publicly accessible and if so, where | 28 | *“The genetic analyses in this study utilize summary-level datasets which are available from the cited sources.”* |
| 20 | **Conflicts of Interest** | All authors should declare all potential conflicts of interest | 28 | *“MCH reports grant support from Genentech, and site principal investigator work, advisory board service for, and in-kind study drug from Novartis, all unrelated to the present work.*  *All other authors have no disclosures to declare.”* |

This checklist is copyrighted by the Equator Network under the Creative Commons Attribution 3.0 Unported (CC BY 3.0) license.

1.	Skrivankova VW, Richmond RC, Woolf BAR, Yarmolinsky J, Davies NM, Swanson SA, et al. Strengthening the Reporting of Observational Studies in Epidemiology using Mendelian Randomization (STROBE-MR) Statement. JAMA. 2021;under review.

2.	Skrivankova VW, Richmond RC, Woolf BAR, Davies NM, Swanson SA, VanderWeele TJ, et al. Strengthening the Reporting of Observational Studies in Epidemiology using Mendelian Randomisation (STROBE-MR): Explanation and Elaboration. BMJ. 2021;375:n2233.
